# Covariation of Zinc Deficiency with COVID-19 Infections and Mortality in European Countries: Is Zinc Deficiency a Risk Factor for COVID-19?

**DOI:** 10.1101/2020.06.12.20105676

**Authors:** Samer Singh

## Abstract

Variables responsible for the differential COVID-19 pandemic severity among countries remain undefined. Zinc, a micronutrient required for immunocompetence, is found deficient in populations. We hypothesized the differential COVID-19 severity observed among European countries could be associated with the Zn-deficiency prevalence. The COVID-19 data from different stages of pandemic, *i.e*., 8 April, 12 and 26 May 2020, were analyzed for covariation with the estimated Zn-deficiency. A significant, relatively stable, but negative correlation of Zn-deficiency with cases per million for the time period [*r*(20): -0.4930 to -0.5335, *p*-value: 0.02720 to 0.0154] and a steady improvement of covariation with deaths per million [*r*(20): -0.4056; *p*-value: 0.0760 on 26 May 2020] was observed. Considering, Zinc’s key immunomodulatory role, widespread deficiency along with the self- and prescribed intervention in different target groups, e.g. children, women, elderly, carefully planned dedicated exploratory studies to understand the basis of the observed association are advisable.

## I. Introduction

The world is currently in the middle of a novel coronavirus, i.e., SARS-CoV-2 caused COVID-19 pandemic. The damage from COVID-19 had been significant in terms of morbidity and mortality. More than 5.67 million people had been infected and over 351 thousand had already died worldwide by 26 May 2020 while more than 1.93 million infections and over 169 thousand deaths were reported from Europe alone (Worldometers, 2020a). The unprecedented severity of COVID-19 in Europe and North America in contrast to its relatively benign course in the countries of Africa, Asia, South America, and Oceania, had been baffling. Several exploratory studies have been initiated based on assumptions as well as our understanding of coronavirus biology to identify and establish protective interventions (WHO, 2020a; WHO, 2020b; Wikipedia, 2020). Recently, a protective correlation between Vitamin D serum levels and the cases and the deaths per million from COVID-19 had been indicated in the European countries supposedly having comparable confounding variables and similar health care infrastructure (Ilie, Stefanescu & Smith, 2020; Singh, Kaur & Singh, 2020). The association had been reassuring given the established role of Vitamin D in overall health and the protection offered from acute respiratory tract diseases through an effect on the host’s renin-angiotensin system comprising ACEs (Cannell, Vieth, Umhau et al, 2006; Xu, Yang, Chen et al, 2017). Identifying such potential protective variables is highly desirable.

Zinc is essential to good health and immunocompetence. Its deficiency is generally associated with a negative impact on overall health, increased susceptibility to disease, and infections (Prasad, 2013; NIH, 2020; EFSA, 2006; Maares & Haase, 2020). In the absence of reliable methods to assess the Zn status of a population, the deficiency or inadequacy in a population is estimated by combining the dietary intake by a population as per FAO’s Food Balance Sheet and the estimated prevalence of stunting (NIH, 2020; EFSA, 2006; Wessells & Brown, 2012; Hess, 2017). Among micronutrients associated with good health, the deficiency of Zinc is quite prevalent ranging from about 5-50 % of the population (Wessells & Brown, 2012). Its high prevalence is associated with increased susceptibility to diseases, mental capacity, complications, high morbidity, and mortality in mothers and neonates (Prasad, 2013; NIH, 2020). Given its role in the overall human health and wellbeing, the countries have tried to overcome the problem through fortification of foods (Prasad, 2013; NIH, 2020; Maares & Haase, 2020; Wessells & Brown, 2012). The countries from the high-income group that consumes a more diverse micronutrient-rich diet are estimated to have Zinc deficiency below 5-10 percent (Wessells & Brown, 2012; Hess, 2017). Incidentally, these had been the countries also worst affected by COVID-19 (Worldometers, 2020a).

Given the global prevalence of Zinc deficiency and its role in human health as well as the immunocompetence (Prasad, 2013; NIH, 2020; EFSA, 2006; Maares & Haase, 2020), we hypothesized that the incidence of COVID-19 in populations (cases) and the adverse outcomes (deaths) would be directly correlated to Zinc deficiency. The analysis of European countries with comparable underlying confounding variables when performed at different stages of the pandemic was envisioned to provide a more robust indication of the correlation as indicated previously (Singh, Kaur & Singh, 2020). The primary aim of the current study was to identify any correlation between the prevalence of Zinc deficiency with the adverse outcomes in the European countries. Additionally, we intended to identify if it affected the number of cases per unit population or crudely the spread of disease. Surprisingly, our analysis of the selected European countries indicated a consistently negative correlation of Zinc deficiency with COVID-19 cases per million populations over the time period and indicated an almost significant correlation with the adverse outcome, i.e., death, which seemed improving with the passage of pandemic.

## II. Material And Methods

The European countries with seemingly comparable confounding variables were considered for the analysis as done previously (Ilie, Stefanescu & Smith, 2020; Singh, Kaur & Singh, 2020). The cases and deaths per million population data for COVID-19 from three time-points, i.e., 8 April, 12 May, and 26 May 2020 (Table 1) were collected from https://www.worldometers.info/coronavirus/ – the coronavirus pandemic data portal. Most of the countries were before COVID-19 infections peak on 8 April 2020 while on 12 and 26 May they were post and late post infections peak stage. The Zinc deficiency estimate data for different countries based on FAO’s food balance datasheet and the prevalence of stunting in the countries were obtained from the previously reported estimate by Wessells & Brown, 2012. All basic statistical analysis of the data, i.e., descriptive, linear regression, calculation of best-fit trend line, Pearson correlation coefficient, and R-squared value (R) was performed in the Microsoft excel as done previously (Singh, Kaur & Singh, 2020).

**Table 1:** COVID-19 pandemic cases and deaths per million population in European Countries with different levels of Zinc deficiency, at three different stages of the current wave of infections.

|  |  | 8 April 2020<br>(Most countries before peak) |  | 12 May 2020<br>(All countries post peak) |  | 26 May 2020<br>(All countries post peak) |  |
| --- | --- | --- | --- | --- | --- | --- | --- |
| Countries | % Zn Deficiency<br>(Sufficiency) | Cases/<br>Million Pop. | Deaths/<br>Million Pop. | Cases/<br>Million Pop. | Deaths/<br>Million Pop. | Cases/<br>Million Pop. | Deaths/<br>Million Pop. |
| Iceland | 3.1(96.9) | 4736 | 18 | 5278 | 29 | 5290 | 29 |
| Norway | 6.2(93.8) | 1123 | 19 | 1500 | 41 | 1547 | 43 |
| Sweden | 6.1(93.9) | 834 | 68 | 2641 | 322 | 3412 | 409 |
| Finland | 4.6(95.4) | 449 | 7 | 1080 | 49 | 1196 | 56 |
| Denmark | 6.2(93.8) | 933 | 38 | 1815 | 92 | 1974 | 97 |
| UK | 4.6(95.4) | 895 | 105 | 3286 | 472 | 3909 | 546 |
| Ireland | 4.0(96.0) | 1230 | 48 | 4685 | 297 | 5015 | 327 |
| Netherlands | 5.0(95.0) | 1199 | 131 | 2497 | 318 | 2661 | 342 |
| Belgium | 6.8(93.2) | 2019 | 193 | 4612 | 751 | 4959 | 806 |
| Germany | 9.0(91.0) | 1309 | 25 | 2060 | 91 | 2164 | 101 |
| France | 3.9(96.1) | 1671 | 167 | 2718 | 408 | 2800 | 437 |
| Switzerland | 4.9(95.1) | 2686 | 103 | 3506 | 213 | 3557 | 221 |
| Italy | 5.8(94.2) | 2306 | 292 | 3636 | 508 | 3813 | 545 |
| Spain | 5.4(94.6) | 3137 | 314 | 5735 | 572 | 6060 | 580 |
| Estonia | 10.5(89.5) | 893 | 18 | 1312 | 46 | 1383 | 49 |
| Czechia | 11.0(89.0) | 488 | 9 | 763 | 26 | 845 | 30 |
| Slovakia | 16.3(83.7) | 125 | 0.4 | 267 | 5 | 277 | 5 |
| Hungary | 8.4(91.6) | 93 | 6 | 340 | 44 | 390 | 52 |
| Turkey | 21.7(78.3) | 453 | 10 | 1657 | 46 | 1884 | 52 |
| Portugal | 6.3(93.7) | 1289 | 37 | 2715 | 112 | 3040 | 132 |
| **Average** | **7.5(92.5)** | **1393.4** | **80.4** | **2605.1** | **222.1** | **2808.8** | **243.0** |
| **STDEV** | **4.5(4.5)** | **1130.0** | **94.6** | **1601.1** | **221.4** | **1681.9** | **238.8** |

**Note:** All values (cases/deaths per million population, Average, STDEV and % Zinc Deficiency in population) rounded off to the displayed decimal places.

## III. Results

A negative correlation between the Zinc deficiency prevalence estimate for the countries and the reported cases of COVID-19 was consistently observed at all three time-points analyzed that covered pre infections peak, i.e.,8 April 2020, and post infections peak, i.e., 12 and 26 May 2020 (see Table 2). The Zinc deficiency of the populations also negatively covaried with adverse outcomes (mortality) per million populations and the correlation seemed to better with the passage of the current wave of COVID-19 infections. Refer to Table 2 below for the comparative estimate of the correlation and Figure 1 for the correlative regression analysis of the data set (R^2^=0.3936 for cases and R^2^=0.2831 for deaths per million) on 26 May 2020. An exploratory estimation of the correlation for COVID-19 affected countries of the world that had seemingly comparable exposure to disease, population structure, and health care response as on 12th May 2020 (> 30,000 cases and upwards of 15,000 tests performed per million of the population) and accounted for 63.7% of total cases, namely, Spain, UK, Russia, Italy, France, Germany, Belgium, Netherlands, Switzerland, USA, Canada, seemed reassuring. For these eleven countries, the Zn deficiency prevalence was exponentially correlated to cases per million with R = 0.3301 and more strongly to deaths per million with R = 0.4307 suggesting a supposedly robust correlation and potential biological significance.

**Figure 1.**
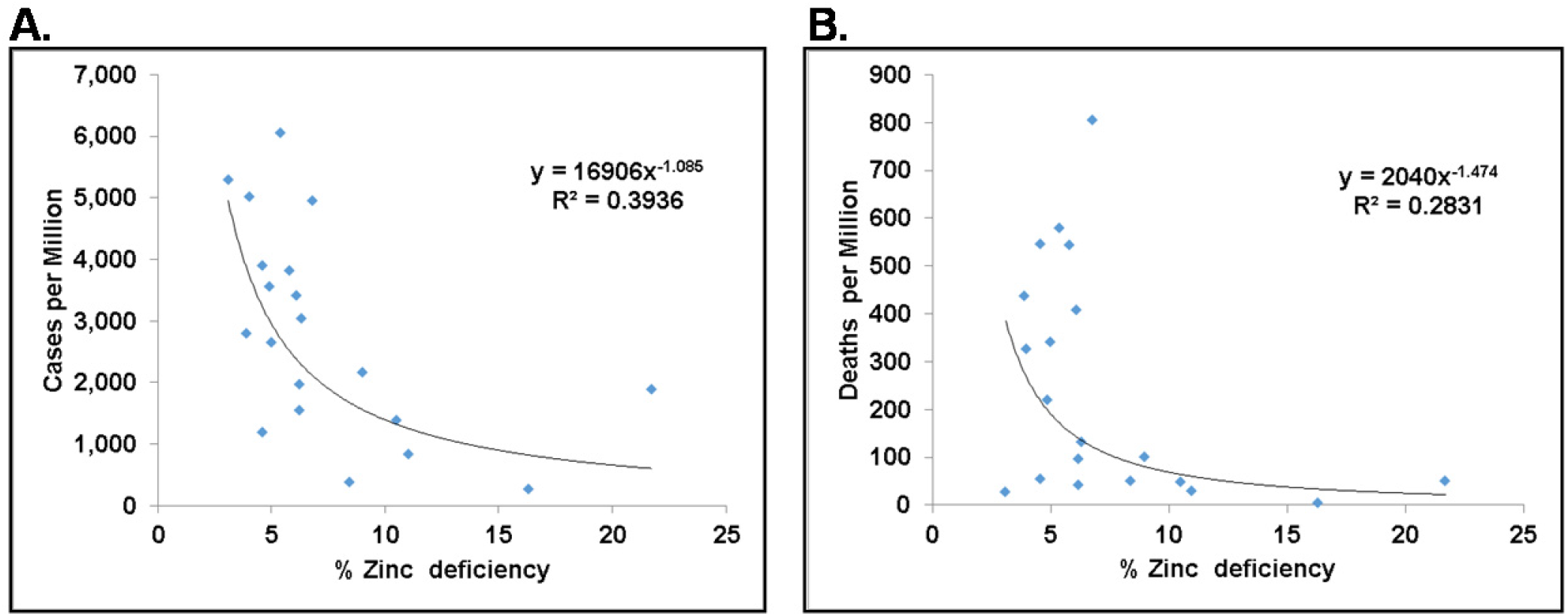
Zinc deficiency in European countries negatively correlated with COVID-19 cases (A) and associated deaths (B) per million population when all countries under study had already passed the peak of current wave of COVID-19 infections (26 May 2020) as per https://www.worldometers.info/coronavirus/.

**Table 2:** Correlation analysis – average Zinc deficiency levels and COVID-19 cases and deaths in select European Countries at different stages of current pandemic

|  | 8 April 2020<br>(Most countries before peak) | 12 May 2020<br>(All countries post peak) | 26 May 2020<br>(All countries post peak) |
| --- | --- | --- | --- |
| Average Cases Per Million Pop. ± STDEV | 1393.4 ± 1130.0 | 2605.2 ± 1601.0 | 2808.8 ± 1681.9 |
| **Correlation: Cases Per Million Pop. vs Zinc deficiency** | $r(20)$: -0.4930;<br>$p$ – value: 0.02720 | $r(20)$: -0.5335;<br>$p$ – value: 0.0154 | $r(20)$: -0.5291;<br>$p$ – value: 0.01644 |
| Average Deaths Per Million Pop. ± STDEV | 80.4 ± 94.6 | 222.1 ± 221.4 | 243.0 ± 238.8 |
| **Correlation: Deaths/M pop. vs Zinc deficiency** | $r(20)$: -0.3652;<br>$p$ – value: 0.1133 | $r(20)$: -0.4027;<br>$p$ – value: 0.0784 | $r(20)$: -0.4056;<br>$p$ – value: 0.0760 |
| % Zinc Deficiency (Sufficiency) ± STDEV | 7.5(92.5) ± 4.5(4.5) | | |

**Note:** The average COVID -19 cases and deaths per million populations for the countries, and the Zinc deficiency (insufficiency) in the population were rounded off to one decimal place. The correlation estimates and p-values were rounded off to four decimal places.

## IV. Discussion

Our analysis identified a negative association between the Zinc deficiency prevalence in the European countries and the COVID-19 impact in terms of cases and deaths per million of population. The observed covariation should be evaluated for its biological significance, if any, by evaluating COVID-19 patient data to allow its potential translation into preventive measures.

In the absence of Zinc storage in the body, its daily intake is required to maintain healthy levels (Prasad, 2013; Maares & Haase, 2020). The recommended daily Zinc intake varies with the age – increasing from 2 mg for newborns to 8 mg for children 9-13 years old. Its recommended intake is 11 mg for >14 years males while 8- 13 mg for females depending upon their physiological status (NIH, 2020) or lower (EFSA, 2006). The countries of Europe and North America have low Zinc deficiency as compared to other parts of the world (Wessells & Brown, 2012; Hess, 2017), rather there exists chances of excessive Zinc intake from food and supplements (NIH, 2020; EFSA, 2006). Considering the associated adverse effect on immunity, lower level of HDL, low copper levels its upper no-observed-adverse-effect level (NOAEL) and tolerable upper intake limit (UL) have been defined by EFSA as daily intake of 50 mg and 25 mg, respectively (EFSA, 2006). For children and adolescents 1-17 years of age, the recommended UL range is 7-22 mg based upon age and body weight. Zinc deficiency is not considered as a concern as for all the age groups the estimated 97.5 percentile of total Zinc intake is closer to the recommended ULs (EFSA, 2006). In the USA, a 20% reduced no adverse effect upper daily limit for Zinc has been suggested, i.e., >40mg for people older than 18 years while for youngers it is from 4-34 mg depending upon the age and body weight (NIH, 2020).

The estimated Zinc deficiency among the European countries, though low, ranged from 3.1 % (Iceland) to 21.7% (Turkey) based upon their dietary preferences (Wessells & Brown, 2012). Countries with a less diverse diet, lower animal protein intake and increased intake of phytate containing diet happen to have a higher prevalence of Zinc deficiency. Generally, the countries with higher Zn deficiency prevalence, e.g., Turkey (21.7%), Slovakia (16.3%), Czechia (11%), Estonia (10.5%), Germany (9%), Hungary (8.4%), Portugal (6.3%), Denmark (6.2%), Norway (6.2%) had reported lesser no of cases per million population as compared to lower Zinc deficiency prevalence countries, e.g., Iceland (3.1), France (3.9), Ireland (4%), UK (4.6%) Switzerland (4.9%) Spain (5.4%), Italy (5.8%). However, the difference in the rate of deaths reported per million population by the above indicated differentially Zinc deficient countries was lesser.

Worldwide estimates indicate lower adverse outcomes from COVID-19 in children, adolescents, and women (Worldometers, 2020b). The Zinc deficiency in children, adolescents, women, vegetarians, and alcoholics is widespread (NIH, 2020; EFSA, 2006). Among the countries of Europe, the children, adolescents, and women seemed to be relatively less affected (Worldometers, 2020b). These are the same constituents of the population that tend to have higher Zinc needs and associated physiological deficiency in the body and have a higher prevalence of Zinc deficiency as compared to the estimated deficiency prevalence in the population (NIH, 2020; EFSA, 2006; Maares & Haase, 2020; Wessells & Brown, 2012; Hess, 2017). Interestingly, women appear to be more protected from COVID-19 than males in European countries (Worldometers, 2020b). The COVID- 19 associated mortality in women is estimated to be about two-fold lower than that in men. Remarkably, women tend to have a higher prevalence of Zinc deficiency deviating about two fold from that estimated for a population as per surveys performed in low and middle-income countries (Hess, 2017). The relationship may be coincidental or causal only later studies can reveal.

Elderly people (>60-65 years) in Europe had been affected most by COVID-19. Although there is insufficient data to reliably estimate the Zinc deficiency in the elderly, it is perceived to be quite prevalent in the elderly with less than 50% of daily recommended intake being common (Mocchegiani, Romeo, Malavolta et al, 2013). Despite the lower Zinc levels in elderly, the altered Zn homeostasis (lower intercellular availability), altered interplay with copper and Iron homeostasis, the supposed immunosenescence and Zinc level alteration as a result of fortification, supplementation, and medication (NIH, 2020; EFSA, 2006; Maares & Haase, 2020) along with other underlying pathologies could be responsible for the relatively higher mortality in the elderly (Worldometers, 2020b). The existence of potential correlations may be established by future epidemiological studies.

## V. Conclusion

A negative correlation between Zinc deficiency and the COVID-19 infections and mortality is observed in European countries. The basis as well as biological significance of such an observation, if any, is unknown. Carefully designed studies investigating the basis of increased vulnerability of target groups and the elderly could reveal the role of Zinc and other key micronutrients such as Cu, Fe and Se in COVID-19 and may help devise better strategies for future COVID-19 control. If the observed correlation is to be believed as having cause and effect relationship, it may be predicted that the inherently or transiently higher Zinc levels resulting from supplementation or medication may be associated with higher COVID-19 infections and mortality in patients. A scrutiny of the COVID-19 patients’ data for potential Zinc status alteration, such as resulting from the intake of vitamin C, organic acids, animal protein, phytate content along with Zinc supplementation and the administration of Zn status altering drugs such as Hydroxychloroquine would shed light on the biological significance of the observed covariation of the prevalence of Zinc deficiency with COVID-19 cases and mortality in different target populations and its potential preventive role. So, more evidence-gathering follow-up exercises for the closer scrutiny of the COVID-19 patients' medical records, including the micronutrient supplementation and food consumption details is advisable to ascertain the role of Zinc or other key micronutrients on COVID-19 pandemic and potentially find an early solution to COVID-19.

## Data Availability

All data is available in the manuscript.

## Author Contributions Statement

SS conceived the idea and written the manuscript.

## Competing Interests Statement

There is no conflict of interest with any person or entity to disclose.

## Ethical Statement

The study complied with the existing ethical standards.

## Acknowledgment

No specific source of funding was utilized for the current study. However, SS acknowledges the funding support from Banaras Hindu University to his laboratory.

## Notes

### Competing Interest Statement

The authors have declared no competing interest.

### Funding Statement

SS acknowledges the funding support from Banaras Hindu University to his laboratory. No external funding specific to this work was received

### Summary of Updates

Author list updated. Minor language corrections at places. Figures revised and included in main file. References updated.

